# The Association Between Estimated Glomerular Filtration Rate and Left Ventricular Function in Children With Chronic Kidney Disease

**DOI:** 10.1101/2024.09.29.24314578

**Authors:** Heshini Dalpathadu, Andrew Wade, Steven Greenway

**Author notes:** These authors contributed equally to this work (Andrew Wade, Steven Greenway). Corresponding author: Steven Greenway MSc, MD, FRCPC, Departments of Cardiac Sciences, Pediatrics, Biochemistry & Molecular Biology Cumming School of Medicine, University of Calgary 3330 Hospital Dr. NW Calgary, Alberta, Canada T2N 1N4.

## Abstract

**Background:** Cardiovascular disease is the leading cause of morbidity and mortality in pediatric patients with chronic kidney disease (CKD). However, the kidney-heart relationship in this population remains poorly understood, particularly in the context of dialysis modality and duration. This study aims to investigate the associations between estimated glomerular filtration rate (eGFR), left ventricular ejection fraction (LVEF), left ventricular (LV) mass, and dialysis modality and duration in pediatric CKD patients.

**Methods:** This retrospective study included 16 pediatric CKD patients (median age 3.6 years; 31.3% female and 68.75% male), stratified by the presence of cardiac dysfunction (LVEF ≤ 50%). Clinical data, including eGFR, LVEF, LV mass, and dialysis history (hemodialysis or peritoneal dialysis), were collected. Independent T-tests, Wilcoxon Two-Sample tests, and Spearman’s correlations were performed to assess renal and cardiac function relationships. Multivariate regression models were employed to evaluate predictors of LVEF over time.

**Results:** Cardiac dysfunction was observed in 25% of the cohort, with significantly lower LVEF and fractional shortening compared to those without dysfunction. Patients with cardiac dysfunction were younger at CKD diagnosis (p < 0.0001), suggesting an earlier progression of renal and cardiac impairment. Following dialysis, eGFR significantly decreased in patients without cardiac dysfunction (p < 0.0001) but remained unchanged in those with dysfunction. Conversely, LVEF improved post-dialysis in patients with cardiac dysfunction (p = 0.0034) but remained stable in those with normal cardiac function. Prolonged dialysis duration was negatively correlated with eGFR (r = –0.31, p = 0.008) and LV mass (r = –0.26, p = 0.024). Hemodialysis duration was positively correlated with LVEF (r = 0.73, p < 0.001), suggesting potential cardiovascular benefits from prolonged hemodialysis treatment.

**Conclusions:** Pediatric CKD patients, particularly those with cardiac dysfunction, experience significant alterations in both renal and cardiac parameters, requiring tailored dialysis strategies in this population.

## INTRODUCTION

In adults and children, CKD is defined as the progressive and irreversible development of abnormalities in kidney structure and function(1,2). CKD is diagnosed based on a glomerular filtration rate (GFR) <60 mL/min/1.73 m^2^, albuminuria >30 mg/24 h, or the presence of other markers of renal dysfunction (e.g. electrolyte or structural abnormalities), which are present for greater than three months(3). The etiology of adult and pediatric CKD significantly differs(4). Unlike adult CKD, which is predominantly secondary to other disease processes (e.g. diabetic nephropathy, hypertension, heart failure), pediatric CKD is associated with congenital anomalies of the kidney and urinary tract (CAKUT), hereditary nephropathies and acquired diseases like glomerulonephritis(4). There is a higher incidence of CV-related complications and mortality among both adult and pediatric CKD patients(1). Cardiovascular (CV) disease is the leading cause of mortality among pediatric patients with CKD(5).

Previous studies have examined the relationship between kidney function and cardiac function; however, these studies were limited to adult or elderly populations (6,7). Given the differences in CKD between adults and children, applying these findings to pediatric patients is both challenging and inaccurate (8). As a result, the kidney-heart relationship in pediatric patients remains poorly understood.

Dialysis, whether through hemodialysis (HD) or peritoneal dialysis (PD), remains a cornerstone of managing advanced CKD in these patients (8). However, there is currently no research that categorizes CKD patients based on their dialysis modality. Additionally, dialysis itself may impose additional cardiovascular stress, potentially leading to adverse outcomes such as left ventricular hypertrophy (LVH) or declines in ejection fraction (8,9). Yet, there is currently no definitive evidence to support these claims, nor is it clear if one dialysis method is more detrimental than the other. As such, kidney transplantation continues to be the only reliable, long-term treatment of CKD in these patients.

In this study, we aim to comprehensively investigate the relationships between estimated glomerular filtration rate (eGFR), left ventricular ejection fraction (LVEF), left ventricular (LV) mass, and dialysis duration in pediatric patients with CKD. Furthermore, we explore the influence of sex on these relationships.

## METHODS

### Study Design and Population

This retrospective study was approved by the Conjoint Health Research Ethics Board of the University of Calgary. We approached and recruited patients between the ages 0-18 with CKD who presented to the Nephrology Clinic at the Alberta Children’s Hospital until May 2022. The inclusion criteria for the study were as follows: patients under 18 years of age at the time of CKD diagnosis; patients with complete longitudinal records of cardiac function, including LVEF and LV mass, renal function measured by eGFR, and treatment history, including duration and modality of dialysis, and/or kidney transplantation during the study period. The patient population included those who had not yet initiated dialysis, had initiated PD or HD and those who had received kidney transplants. Written informed consent was obtained from all patients. All data were anonymized prior to analysis.

### Data Collection

Patient records were reviewed to extract key clinical parameters. LVEF and LV mass were measured using echocardiography, with values recorded at multiple time points throughout the disease course. Left ventricular dimensions were measured with M-mode and LVEF was calculated by using the Teichholz method. The presence of cardiac dysfunction was defined as an LVEF of 50% or lower in accordance with established clinical guidelines. Renal function was assessed by eGFR, calculated using the creatinine-based Schwartz formula, which is widely used in pediatric populations. Dialysis history was documented, along with any transitions to kidney transplantation. Demographic information, including patient age, sex, and baseline clinical characteristics, was collected.

### Statistical Analysis

The cohort was stratified based on the presence or absence of cardiac dysfunction (LVEF ≤ 50%). Data were evaluated using GraphPad Prism version 10.3.1(464). Comparisons between two independent groups were performed using the Independent T-Test for normally distributed continuous variables, while the Wilcoxon Two-Sample Test (Mann-Whitney U Test) was applied to continuous variables with non-normal distributions. Fisher’s Exact Test was used to analyze categorical variables. Correlations between estimated GFR and echocardiographic parameters were assessed by Spearman’s correlation test. A p-value of <0.05 was considered statistically significant.

Linear regression analyses were used to assess the cross-sectional associations of eGFR (independent variable) with LVEF and LV mass (dependent variables in separate models). A multivariate regression model was employed to identify predictors of LVEF change over time. Variables included in the model were baseline LVEF, LV Mass, eGFR, dialysis duration, and sex. Interaction terms were included to explore potential synergistic effects between variables.

## RESULTS

### Study Population

A total of 16 pediatric patients with CKD were included in this study (Table 1). The median age at the time of CKD diagnosis was 3.6 (0.8, 6.1) years, with 68.75% being male and 31.25% female. Among the total cohort, 10 underwent PD, 5 patients underwent HD, and 3 patients received a kidney transplant during the study period. 5 patients did not undergo either type of dialysis. Cardiac dysfunction (LVEF ≤ 50%) was observed in 25% of the patients.

**Table 1:** Patient Demographics and characteristics of the 16 pediatric patients stratified according to the presence of cardiac dysfunction (defined as a left ventricular ejection fraction of less than 50%).

| Variable | All Subjects<br>(n=16) | Patients with<br>Cardiac<br>Dysfunction (n=4) | Patients without<br>Cardiac<br>Dysfunction (n=12) | P-Value<br>Independent T-Test<br>*Wilcoxon Two-Sample Test*<br>(Fisher's Exact Test) |
| --- | --- | --- | --- | --- |
| Age | 3.6 (0.8, 6.1) | 0.4 (0.1, 3.4) | 3.9 (1.9, 7.4) | <0.0001 **** |
| Sex (Female) | 5 (31.3%) | 2 (50.0%) | 3 (25.0%) | (0.5467) |
| eGFR (mL/min/1.73 m<sup>2</sup>) | 14.2 (9.2, 24.4) | 18.7 (11.8, 28.6) | 11.7 (8.9, 20.3) | 0.0983 |
| LVEF (%) | 63.6 ± 13.6 | 41.9 ± 5.0 | 70.1 ± 6.8 | <0.0001 **** |
| LV Mass (C) dI (g/m<sup>2</sup>) | 81.9 (60.6, 123.6) | 93.9 (75.7, 128.0) | 76.0 (53.0, 122.3) | 0.1109 |
| Fractional Shortening (%) | 36.19 ± 8.4 | 27.3 ± 10.3 | 38.8 ± 5.6 | <0.0001 **** |
| Peritoneal Dialysis | 10 (62.5%) | 3 (75.0%) | 7 (58.33%) | (>0.9999) |
| Hemodialysis | 5 (31.3%) | 2 (50.0%) | 3 (25.0%) | (0.5467) |
| Any Dialysis | 12 (75.0%) | 3 (75.0%) | 9 (75.0%) | (>0.9999) |
| Transplant | 3 (18.8%) | 2 (50.0%) | 1 (8.3%) | (0.1357) |

Data are presented as mean ± SD or median (25th, 75th percentiles) for continuous variables and number (%) for categorical variables. Abbreviations: eGFR: estimated glomerular filtration rate; LV: left ventricular; LVEF: left ventricular ejection fraction.

When the study participants were stratified based on their cardiac function (Table 1), patients with cardiac dysfunction had a significantly younger median age compared to those without cardiac dysfunction (p < 0.0001). There were no significant differences in eGFR values or sex distribution between the two groups. As expected, patients with cardiac dysfunction had significantly lower LVEF (p < 0.0001), confirming the presence of systolic dysfunction in this group. In addition, these patients had significantly lower fractional shortening (p < 0.0001), another marker of systolic function. LV mass was higher in the cardiac dysfunction group, though this difference was not statistically significant (p = 0.1109). No significant differences were observed between the two groups regarding the need for PD (p > 0.9999), HD (p = 0.5467), any form of dialysis (p > 0.9999) or for a kidney transplant (p = 0.1357).

### Renal and Cardiac Function Before and After Dialysis

Table 2 summarizes key renal and cardiac parameters before and after dialysis in pediatric patients with CKD, stratified by the presence or absence of cardiac dysfunction. The p-values reflect the comparisons between four key conditions: before and after dialysis in patients with cardiac dysfunction ([A] vs [B]), before and after dialysis in patients without cardiac dysfunction ([C] vs [D]), and across groups both before ([A] vs [C]) and after dialysis ([B] vs [D]).

**Table 2:** Comparative analysis of renal and cardiac parameters before and after dialysis in pediatric patients.

| Variables | CKD with Cardiac Dysfunction |  | CKD without Cardiac Dysfunction |  | P-Value Independent T-Test *Wilcoxon Two-Sample Test* | P-Value Independent T-Test *Wilcoxon Two-Sample Test* | P-Value Independent T-Test *Wilcoxon Two-Sample Test* | P-Value Independent T-Test *Wilcoxon Two-Sample Test* |
| --- | --- | --- | --- | --- | --- | --- | --- | --- |
|  | Before dialysis [A] | After dialysis [B] | Before dialysis [C] | After dialysis [D] | (Before vs After Dialysis in CKD with Cardiac Dysfunction) [A vs B] | (Before vs After Dialysis in CKD without Cardiac Dysfunction) [C vs D] | (Before Dialysis in CKD with vs without Cardiac Dysfunction) [A vs C] | (After Dialysis in CKD with vs without Cardiac Dysfunction) [B vs D] |
| eGFR | 22.4 (13.2, 28.1) | 13.0 (7.1, 19.3) | 19.5 (11.7, 35.3) | 10.0 (7.2, 14.3) | 0.0780 | <0.0001 **** | 0.7737 | 0.0812 |
| LVEF (%) | 46.0 ± 6.8 | 62.6 ± 14.0 | 69.7 ± 7.2 | 70.4 ± 7.0 | 0.0034 ** | 0.5736 | <0.0001 **** | 0.0039 ** |
| LV mass (C) dI (g/m2) | 130.2 (40.8, 136.4) | 70.0 (57.8, 90.0) | 77.1 (52.3, 138.5) | 75.7 (58.1, 104.4) | 0.2338 | 0.6777 | 0.7285 | 0.3498 |
| Fractional Shortening (%) | 29.1 ± 11.74 | 35.5 ± 8.5 | 38.0 ± 5.7 | 39.4 ± 5.5 | 0.0831 | 0.3632 | 0.0100 * | 0.0211 * |

Data are presented as mean ± SD or median (25th, 75th percentiles). Abbreviations: CKD: chronic kidney disease; eGFR: estimated glomerular filtration rate; LV: left ventricular; LVEF: left ventricular ejection fraction.

Despite eGFR declining after dialysis in patients with cardiac dysfunction, the decrease was not statistically significant. In contrast, in patients without cardiac dysfunction, eGFR significantly decreased after dialysis (p < 0.0001). When comparing eGFR between patients with and without cardiac dysfunction, eGFR was not significantly different both before dialysis and after dialysis.

In patients with cardiac dysfunction, LVEF significantly increased after dialysis (p = 0.0034), indicating a marked improvement in systolic function. In contrast, no significant change in LVEF was observed before and after dialysis in patients with normal cardiac function (p = 0.5736). However, LVEF was significantly lower in patients with cardiac dysfunction compared to those without, both before (p < 0.0001) and after dialysis (p = 0.0039).

Although LV mass decreased after dialysis in patients with cardiac dysfunction, it remained relatively stable in patients without cardiac dysfunction. However, neither comparison was statistically significant. Similarly, no significant differences in LV mass were found between the two cardiac function groups before dialysis or after dialysis.

Fractional shortening showed a trend toward improvement in patients with cardiac dysfunction after dialysis, though this did not reach significance. In patients without cardiac dysfunction, fractional shortening remained relatively stable. When comparing between groups, fractional shortening was significantly lower in patients with cardiac dysfunction before dialysis (p = 0.0100) and after dialysis (p = 0.0271).

### Correlation Between eGFR and LVEF

Table 3 presents the correlation between eGFR and LVEF, stratified by the presence of cardiac dysfunction in pediatric CKD patients. In the overall population (r = –0.20, 95% CI: –0.41 to 0.04) and in patients without cardiac dysfunction (r = –0.14, 95% CI: –0.39 to 0.14), weak negative correlations were found between eGFR and LVEF, with no statistical significance. Among patients with cardiac dysfunction, a moderate positive correlation was observed between eGFR and LVEF (r = 0.35, 95% CI: –0.18 to 0.72), though this correlation did not reach statistical significance (p = 0.173). This suggests a potential relationship where impaired kidney function (lower eGFR) might be associated with reduced cardiac function (lower LVEF) in this subgroup.

**Table 3:** Correlations between eGFR and LVEF, stratified by cardiac dysfunction.

| Correlation | Correlation Coefficient (95% CI) | P-Value |
| --- | --- | --- |
| eGFR vs. LVEF (Overall) | -0.20 (-0.41, 0.04) | 0.089 |
| eGFR vs. LVEF (Cardiac Dysfunction) | 0.35 (-0.18, 0.72) | 0.173 |
| eGFR vs. LVEF (No Cardiac Dysfunction) | -0.14 (-0.39, 0.14) | 0.312 |

Data are presented as correlation coefficients (95% confidence intervals), with p-values reflecting the statistical significance of the correlations.

### Correlation Between eGFR and LV Mass

Table 4 shows the correlations between eGFR and LV mass across different subgroups. Overall, there was a very weak positive correlation between eGFR and LV mass (r = 0.11, 95% CI: –0.13 to 0.34), though not statistically significant. In patients with cardiac dysfunction, a weak positive correlation was observed (r = 0.28, 95% CI: –0.24 to 0.68), but also lacked significance. In those without cardiac dysfunction, the correlation was negligible (r = 0.02, 95% CI: –0.25 to 0.29) and not significant.

**Table 4:** Correlations between eGFR and LV mass, stratified by cardiac dysfunction.

| Correlation | Correlation Coefficient (95% CI) | P-Value |
| --- | --- | --- |
| eGFR vs. LV Mass (Overall) | 0.11 (-0.13, 0.34) | 0.350 |
| eGFR vs. LV Mass (Cardiac Dysfunction) | 0.28 (-0.24, 0.68) | 0.266 |
| eGFR vs. LV Mass (No Cardiac Dysfunction) | 0.02 (-0.25, 0.29) | 0.871 |

Data are presented as correlation coefficients (95% confidence intervals), with p-values reflecting the statistical significance of the correlations.

### Correlation Between eGFR and LVEF Stratified by Sex

Correlations between eGFR and LVEF, stratified by sex and cardiac function status, were analyzed (Table 5). Among females, a weak negative correlation was observed overall (r = –0.32, 95% CI: –0.72 to 0.24). In female children with cardiac dysfunction, there was a moderate positive correlation (r = 0.57), while a negative correlation (r = –0.39) was noted in those without cardiac dysfunction.

**Table 5:** Correlations between eGFR and LVEF, stratified by sex.

| Correlation | Correlation Coefficient (95% CI) | P-Value |
| --- | --- | --- |
| eGFR vs. LVEF (Overall; Females) | -0.32 (-0.72, 0.24) | 0.242 |
| eGFR vs. LVEF (Cardiac Dysfunction; Females) | 0.57 | 0.151 |
| eGFR vs. LVEF (No Cardiac Dysfunction; Females) | -0.39 | 0.396 |
| eGFR vs. LVEF (Overall; Males) | -0.10 (-0.36, 0.16) | 0.433 |
| eGFR vs. LVEF (Cardiac Dysfunction; Males) | 0.25 | 0.511 |
| eGFR vs. LVEF (No Cardiac Dysfunction; Males) | -0.07 (-0.34, 0.23) | 0.654 |

Data are presented as correlation coefficients (95% confidence intervals), with p-values reflecting the statistical significance of the correlations.

In the overall male patient population, a very weak negative correlation between eGFR and LVEF was found (r = –0.10, 95% CI: –0.36 to 0.16). Similar to females, male children with cardiac dysfunction showed a positive correlation (r = 0.25), though it was weak. In males without cardiac dysfunction, a negligible negative correlation (r = –0.07) was observed. None of the sex-based correlations between eGFR and LVEF were statistically significant.

### Dialysis Modality Comparisons

Table 6 summarizes the comparison of eGFR and LVEF before and after dialysis in pediatric patients undergoing hemodialysis or peritoneal dialysis. For patients on HD, there were no significant changes in either eGFR or LVEF following treatment. In contrast, the initiation of PD resulted in a statistically significant decline in eGFR (p < 0.0001), reflecting a marked decrease in renal function. Although LVEF showed an increase after PD, this improvement did not reach statistical significance.

**Table 6:**
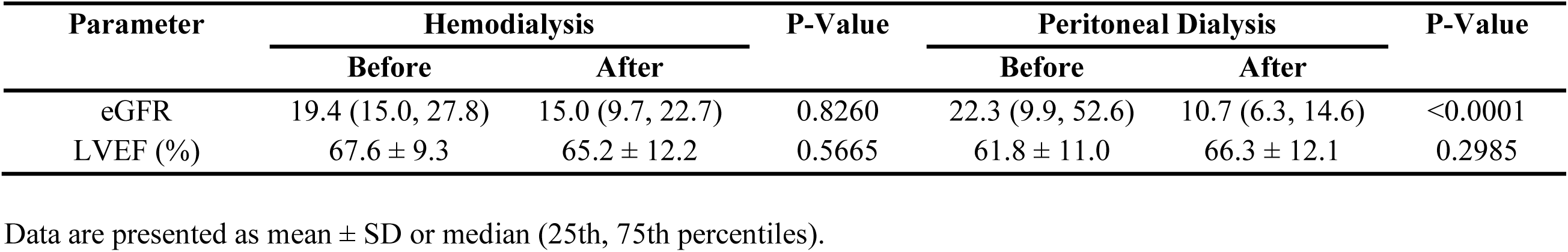
Comparison of eGFR and LVEF Before and After Undergoing Hemodialysis and Peritoneal Dialysis in Pediatric Patients.

| Parameter | Hemodialysis |  | P-Value | Peritoneal Dialysis |  | P-Value |
| --- | --- | --- | --- | --- | --- | --- |
|  | Before | After |  | Before | After |  |
| eGFR | 19.4 (15.0, 27.8) | 15.0 (9.7, 22.7) | 0.8260 | 22.3 (9.9, 52.6) | 10.7 (6.3, 14.6) | <0.0001 |
| LVEF (%) | 67.6 ± 9.3 | 65.2 ± 12.2 | 0.5665 | 61.8 ± 11.0 | 66.3 ± 12.1 | 0.2985 |

Data are presented as mean ± SD or median (25th, 75th percentiles).

The correlations between dialysis duration and key renal and cardiac parameters are shown in Table 7. Overall, a significant moderate negative correlation was observed between dialysis duration and eGFR (r = –0.31, 95% CI: –0.51 to –0.08, p = 0.008), indicating a decline in kidney function with longer dialysis duration. This trend was also significant in patients without cardiac dysfunction (r = –0.30, 95% CI: –0.53 to –0.04, p = 0.021). However, no significant correlation was found between dialysis duration and eGFR in patients with cardiac dysfunction. Interestingly, the duration of PD showed a stronger, albeit non-significant, negative correlation with eGFR compared to HD.

**Table 7:** Correlations between dialysis duration, eGFR, LVEF, and LV mass, stratified by overall population and cardiac dysfunction status. Additional comparisons are shown for peritoneal dialysis (PD) and hemodialysis (HD) durations in relation to renal and cardiac parameters.

| Correlation | Correlation Coefficient (95% CI) | P-Value |
| --- | --- | --- |
| Dialysis Duration vs. eGFR (Overall) | -0.31 (-0.51, -0.08) | 0.008 ** |
| Dialysis Duration vs. eGFR (Cardiac Dysfunction) | -0.30 (-0.69, 0.22) | 0.234 |
| Dialysis Duration vs. eGFR (No Cardiac Dysfunction) | -0.30 (-0.53, -0.04) | 0.021 * |
| PD Duration vs. eGFR (Overall) | -0.43 (-0.79, 0.14) | 0.124 |
| HD Duration vs. eGFR (Overall) | -0.23 (-0.61, 0.24) | 0.312 |
| Dialysis Duration vs. LVEF (Overall) | 0.05 (-0.19, 0.28) | 0.701 |
| Dialysis Duration vs. LVEF (Cardiac Dysfunction) | -0.13 (-0.58, 0.39) | 0.620 |
| Dialysis Duration vs. LVEF (No Cardiac Dysfunction) | 0.05 (-0.22, 0.32) | 0.702 |
| PD Duration vs. LVEF (Overall) | -0.21 (-0.67, 0.38) | 0.475 |
| HD Duration vs. LVEF (Overall) | 0.73 (0.43, 0.89) | <0.001 *** |
| Dialysis Duration vs. LV mass (Overall) | -0.26 (-0.47, -0.03) | 0.024 * |
| Dialysis Duration vs. LV mass (Cardiac Dysfunction) | -0.44 (-0.76, 0.07) | 0.081 |
| Dialysis Duration vs. LV mass (No Cardiac Dysfunction) | -0.21 (-0.45, 0.06) | 0.120 |
| PD Duration vs. LV mass (Overall) | 0.31 (-0.28, 0.73) | 0.274 |
| HD Duration vs. LV mass (Overall) | -0.06 (-0.48, 0.38) | 0.778 |

Data are presented as correlation coefficients (95% confidence intervals), with p-values reflecting the statistical significance of the correlations.

When examining the relationship between dialysis duration and LVEF, no significant correlations were found either overall or in subgroups. Notably, however, a significant strong positive correlation was identified between HD duration and LVEF (r = 0.73, 95% CI: 0.43 to 0.89, p < 0.001), suggesting that longer HD treatment may be associated with improved cardiac function.

With regard to LV mass, a significant weak negative correlation was found between dialysis duration and LV mass overall (r = –0.26, 95% CI: –0.47 to –0.03, p = 0.024), indicating that prolonged dialysis was associated with lower LV mass. Although this trend was more pronounced in patients with cardiac dysfunction (r = –0.44, 95% CI: –0.76 to 0.07, p = 0.081), it did not reach statistical significance. No significant correlations were identified between PD or HD duration and LV mass.

### Multivariate Regression Analysis

The results of the multivariate regression analysis, as shown in Table 8, evaluate the predictors of LVEF in pediatric patients, stratified by cardiac dysfunction status. Across all models, LV mass, dialysis duration, and their interaction with LVEF were examined.

**Table 8:** Multivariate Regression Analysis. Predictors of LVEF in pediatric patients, stratified by overall population, patients with cardiac dysfunction, and patients without cardiac dysfunction. Regression models include LV mass, dialysis duration, and their interaction as independent variables.

| Predictor | Regression Coefficient (95% CI) | Standard Error | P-Value |
| --- | --- | --- | --- |
| Intercept (Overall) | 68.32 (59.53, 77.11) | 4.41 | <0.0001 |
| LV Mass (Overall) | -0.07 (-0.16, 0.02) | 0.04 | 0.1084 |
| Dialysis Duration (Overall) | -0.003 (-0.03, 0.02) | 0.01 | 0.7878 |
| Interaction (EF*Dialysis Duration) (Overall) | 0.0002 (-0.0001, 0.0005) | 0.0002 | 0.3143 |
| Intercept (Cardiac Dysfunction) | 46.22 (33.36, 59.09) | 5.96 | <0.0001 |
| LV Mass (Cardiac Dysfunction) | -0.04 (-0.15, 0.08) | 0.05 | 0.5181 |
| Dialysis Duration (Cardiac Dysfunction) | 0.03 (-0.09, 0.16) | 0.06 | 0.5824 |
| Interaction (EF*Dialysis Duration) (Cardiac Dysfunction) | -0.0006 (-0.002, 0.001) | 0.0007 | 0.4254 |
| Intercept (No Cardiac Dysfunction) | 73.37 (68.56, 78.18) | 2.397 | <0.0001 |
| LV Mass (No Cardiac Dysfunction) | -0.04 (-0.088 to 0.007) | 0.02 | 0.0945 |
| Dialysis Duration (No Cardiac Dysfunction) | -0.007 (-0.019, 0.005) | 0.006 | 0.2621 |
| Interaction (EF*Dialysis Duration) (No Cardiac Dysfunction) | 0.0001 (-5.076e-005, 0.0003) | 7.780e-005 | 0.1817 |

Data are presented as correlation coefficients (95% confidence intervals), with p-values reflecting the statistical significance of the correlations.

In the overall population, the intercept for the model was 68.32 (95% CI: 59.53, 77.11; p < 0.0001), indicating the baseline LVEF when LV mass and dialysis duration are at zero. LV mass showed a very weak negative association with LVEF, though this was not statistically significant. Dialysis duration and its interaction with LV mass did not significantly affect LVEF.

In patients with cardiac dysfunction, the intercept was 46.22 (95% CI: 33.36, 59.09; p < 0.0001), reflecting a lower baseline LVEF in this subgroup. There were no significant associations observed between LV mass, dialysis duration, or their interaction and LVEF. In contrast, in patients without cardiac dysfunction, the intercept was 73.37 (95% CI: 68.56, 78.18; p < 0.0001), suggesting a higher baseline LVEF. LV mass exhibited a near-significant negative association with LVEF (p = 0.0945), while dialysis duration and its interaction with LV mass were not significant predictors.

## DISCUSSION

The intricate interplay between cardiac and renal dysfunction in pediatric patients with CKD presents a formidable challenge for clinicians. This study aimed to explore the complexities of the relationship between cardiac parameters, including LVEF, LV mass, and fractional shortening, and renal parameters, such as eGFR, while also examining the influence of dialysis duration and modality. Our findings provide valuable insights into the heart-kidney axis in this vulnerable patient population and underscore the need for an individualized approach to management, especially in those with concurrent cardiac dysfunction.

One of the key observations from this study is that children with CKD and cardiac dysfunction tend to present at a significantly younger age compared to those without cardiac dysfunction, suggesting that the onset of cardiac abnormalities may be linked to an earlier progression of renal impairment, which may be especially relevant in pediatric patients with CAKUT (10). The early onset of CKD in CAKUT patients, combined with the developmental impact of congenital anomalies, places this population at heightened risk for cardiovascular complications (11). This underscores the importance of early surveillance and intervention in children with CKD secondary to CAKUT, as they may require more aggressive management to mitigate the progression of both renal and cardiac disease.

Our results also highlight the significant burden of systolic dysfunction in children with cardiac abnormalities, as evidenced by lower LVEF and fractional shortening compared to those without cardiac dysfunction. This is consistent with existing literature indicating that CKD contributes to a high burden of cardiovascular disease in pediatric populations, primarily through left ventricular hypertrophy and subsequent systolic dysfunction (9,12).

The analysis of cardiac and renal parameters before and after dialysis revealed important differences between patients with and without cardiac dysfunction. In patients without cardiac dysfunction, eGFR significantly declined after dialysis, reflecting the expected progression of renal disease in this population. In contrast, patients with cardiac dysfunction showed no statistically significant change in eGFR post-dialysis, but they did exhibit a significant improvement in LVEF. This suggests that dialysis may alleviate some of the cardiac stress, potentially by reducing fluid overload and uremic toxin burden, which are known contributors to cardiac dysfunction (13). Conversely, LVEF remained stable in patients without cardiac dysfunction before and after dialysis, indicating that the protective effect of dialysis on cardiac function may be more pronounced in patients with pre-existing cardiac impairments.

Notably, LV mass did not change significantly after dialysis in either group, and there were no significant differences in LV mass between the groups before or after dialysis. This suggests that while dialysis may have an acute effect on cardiac function, particularly in those with dysfunction, its impact on cardiac structure, such as left ventricular hypertrophy, may require longer periods of renal replacement therapy or additional interventions to manifest measurable changes.

Our correlation analyses further emphasize the complexity of the heart-kidney axis in pediatric CKD. While no significant correlations were observed between eGFR and LVEF in the overall population, there was a moderate positive correlation in patients with cardiac dysfunction, suggesting a potential interplay between worsening kidney function and declining cardiac function. This reinforces that while renal function decline is expected in CKD, the direct impact on cardiac function may be more nuanced.

The relationship between dialysis duration and clinical parameters offers another layer of complexity. The significant negative correlation between dialysis duration and eGFR across the overall population suggests that prolonged dialysis may contribute to progressive renal function decline, a well-documented outcome in pediatric CKD. However, the strong positive relationship between HD duration and LVEF indicates potential cardiovascular benefits from prolonged HD treatment. This finding warrants further investigation, as it could reshape our understanding of dialysis-induced cardiac adaptations in pediatric patients potentially by better managing fluid overload and reducing afterload (14). In contrast, the significant negative correlation between dialysis duration and LV mass suggests that while dialysis is life-saving and essential for managing CKD, prolonged exposure may contribute to adverse cardiac remodeling (12). The reduction in LV mass over time could be a double-edged sword: on one hand, it might indicate a reversal of hypertrophy; on the other, it could signal a loss of cardiac muscle mass, potentially heightening the risk of heart failure. Therefore, clinicians should consider these risks when selecting a dialysis modality, particularly for pediatric patients with diagnosed cardiac dysfunction.

In our multivariate regression analysis, neither LV mass nor dialysis duration emerged as significant predictors of LVEF across the cohort. These findings underscore the complexity of predicting cardiac outcomes in pediatric CKD, as multiple factors, including dialysis modality, volume status, uremic toxin levels, and the underlying etiology of CKD, likely contribute to the variability in cardiac response to renal impairment and dialysis (8).

This study is one of the few to comprehensively analyze the cardiorenal interactions in a pediatric CKD population, providing valuable insights into how these kidney and heart clinical parameters differ based on cardiac function status, and dialysis modality. Our findings underscore the critical need for identifying modifiable risk factors and implementing individualized treatment strategies in pediatric CKD patients. Early and proactive management, including timely initiation of renal replacement therapy, could be pivotal in preserving cardiac function and preventing long-term deterioration.

## FUTURE DIRECTIONS

The findings of this study open several avenues for future research. Longitudinal studies with larger, diverse cohorts, extended follow-up and more comprehensive data collection are needed to confirm and expand upon our findings. Specifically, the role of sex in modulating the relationship between renal and cardiac function in pediatric CKD patients deserves further investigation, as does the long-term impact of different dialysis modalities on cardiac outcomes. Additionally, exploring the genetic and molecular mechanisms underlying the observed variability in patient responses could lead to the development of more targeted therapies. Finally, integrating these findings into clinical practice will require the development of new guidelines that consider the complex cardiorenal interactions in pediatric CKD patients.

## CONCLUSION

In conclusion, this study highlights the complex and multifaceted relationship between cardiac and renal dysfunction in pediatric CKD patients. By shedding light on these interactions, we hope to pave the way for more effective management strategies that improve the long-term outcomes for these vulnerable patients.

## ACKNOWLEDGEMENTS

The authors would like to gratefully acknowledge the efforts of the Mozell Family Analysis Core for their invaluable guidance with the biostatistical analysis of this project. We would also like to extend our thanks to Stephanie Hunter and Carlie Barath, nurses at the Nephrology Clinic at Alberta Children’s Hospital, for their assistance in data acquisition.

## Data Availability Statement

Data available on request from the authors.

